# A Multi-Center Randomized, Double-Blind, Placebo Controlled, Parallel Group, Phase IIa Study to Evaluate the Efficacy, Safety and Tolerability of an Anthocyanin Rich Extract (ACRE) in Patients with Ulcerative Colitis

**DOI:** 10.1101/2024.07.19.24310589

**Authors:** Luc Biedermann, Michael Doulberis, Philipp Schreiner, Ole Haagen Nielsen, Frans Olivier The, Stephan Brand, Sabine Burk, Petr Hruz, Pascal Juillerat, Claudia Krieger-Grübel, Kristin Leu, Gabriel Leventhal, Benjamin Misselwitz, Sylvie Scharl, Alain Schoepfer, Frank Seibold, Hans Herfarth, Gerhard Rogler

## Abstract

**Background:** In an open label pilot study dried bilberries were effective in inducing clinical, endoscopic and biochemical improvement in ulcerative colitis (UC) patients. Aim was the investigation of efficacy of anthocyanin rich extract (ACRE), the presumptive active ingredient of bilberries, in a controlled clinical trial in moderate-severe UC.

**Methods:** We performed a multicenter randomized, placebo-controlled, double-blind study (planned initially for 100 patients; premature termination due to COVID-19 pandemic). Patients had moderate-severe active UC at screening (Mayo-score 6-12, endoscopic sub-score at least 2) and were randomized at baseline (verum: placebo, 2:1). Continuation of all UC-directed stable medical therapy was allowed. Primary endpoint was clinical response at week 8 (reduction of total Mayo-score at least 3 points). Biochemical (fecal calprotectin) and centrally-read endoscopic response were amongst the secondary endpoints.

**Results:** Out of 48 patients screened in six Swiss trial centers, 34 were randomized. Eighteen ACRE and eight placebo patients could be analyzed in the Per-Protocol-Set. Half (9/18) of ACRE patients and 3/8 of placebo patients revealed clinical response at week 8 (CI 0.399-6.963; p=0.278). An improvement of the Mayo-score was observed in 77.8% of ACRE treated patients (62.5% of placebo). Fecal calprotectin dropped from 1049+/-1139 to 557+/-756μg/g feces in the ACRE but not in the placebo group (947+/-1039 to 1040+/-1179; p=0.035). Adverse events were rare.

**Conclusions:** ACRE therapy was not significantly superior to placebo at inducing a clinical response. However, placebo response was unusual high. Moreover, there was a significant calprotectin decrease at end of treatment, indicative of ACRE biochemical efficacy in UC.

**Study Highlights:** *What is known:*

- Dried bilberries have been reported to ameliorate active ulcerative colitis (UC) in an uncontrolled pilot trial
- Anthocyanins (flavonoids) are regarded to be the active anti-inflammatory compound of bilberries
- An anthocyanin rich extract (ACRE) of bilberries was reported to ameliorate colitis in mouse models

*What is new here:*

- In a multi-center randomized, double-blind, placebo controlled, parallel group study in patients with moderate to severe active UC, ACRE did not reach the statistical endpoint of clinical response
- An unusually high placebo response was observed
- ACRE induced significant biochemical response with significant decrease in calprotectin levels

## Introduction

Ulcerative colitis (UC) comprises along with Crohn’s disease, the most common forms of inflammatory bowel diseases (IBD). UC is an idiopathic, chronic relapsing pathology of the colon affecting mainly young adults with a peak age range between 30 - 40 years and no gender predominance. Amongst the most typical symptoms are severe, often bloody diarrhea, as well as abdominal discomfort. Although the clinical course is variable, most patients suffer from recurrent flares and an unpredictable disease course.^1,2^ Quality of life may be severely impaired by persistent, frequent, recurrent or waxing and waning symptoms.^3^

Despite the incurable nature of the disease, about two thirds of all UC patients with moderate disease activity can be successfully treated with mesalamine (5-ASA).^4,5^ Nevertheless, patients who do not achieve a sufficient response remain a clinical challenge and up to 10% of all patients have to undergo colectomy in the long-term, according to a very recent (2023) systematic review and meta-analysis.^6^

Although there is a plethora of potential effective drugs to treat UC, the overall profile of those is far from being ideal: They harbor a considerable risk of short- and long-term toxicity and numerous side effects.^7–9^ Moreover, the annual costs of newer treatment options (such as biologics and small molecules) are often high.^10,11^ Most importantly, even the latest treatments demonstrate efficiency in only a proportion of UC patients.^12^ Therefore, the development of further medical treatment options, with favorable cost-benefit ratios and salutary side-effect profiles clearly represents an urgent requirement for UC management. In this respect, herbal and natural compound treatments constitute an appealing therapy option for UC patients. Secondly, many Swiss patients long for more natural therapies. In this context, several relevant studies report positive effect of curcuma^13^ (*Curcuma zedoaria,* an Indian spice), or of boswellia tree resin (*Boswellia serrata*) as well as aloe vera^14,15^ on UC patients.

In recent years there has been a rising interest in natural flavonoids, a subgroup of polyphenols, due to their beneficial effects on many conditions including cardiovascular disease and cancer. Anthocyanins (AC), compounds belonging to flavonoids, are abundant in red, blue and black berries, but also present in red wine and dark colored vegetables.^16,17^

AC have been associated with many protective biological effects, including antioxidative, anticarcinogenic, antimicrobial, and anti-inflammatory properties.^15,18,19^ Due to their phenolic structure, AC exhibit anti-oxidative capacity *in vivo* as they scavenge reactive oxygen species (ROS),^18,20^ a classical effect of 5-ASA.^21^ AC interrupt pro-inflammatory signaling and are inhibitors of 5-lipoxygenase, a key enzyme implicated in the arachidonic acid pathway, for the biosynthesis of active leukotrienes (mainly via the unstable intermediate LTA_4_ to LTB_4_ and 5-HETE^22,23^). In the presence of flavonoids monocytes release less tumor necrosis factor (TNF) and interleukin (IL) – 8.^24^ AC also impede activation of nuclear factor κB (NF-ĸB) by inhibition of proteasomal function. NF-ĸB as well as TNF and IL-8 constitute key molecules orchestrating inflammation in IBD.^15,25,26^

Several research groups^27–33^ including ours^19,34–39^ reported a beneficial effect of AC on preclinical models of UC. In view of the aforementioned results, the potential of AC in a small uncontrolled pilot trial in 13 patients with UC was tested.^40^ During six weeks, patients received a daily AC-rich extract from bilberries (*Vaccinium myrtillus*). Strikingly, clinical disease activity, as well endoscopic histological and biochemical indicators for intestinal inflammation markedly improved. Side effects were not observed. These data suggest AC as a potential adjunctive treatment option in UC with very few, if any, side effects.^40,41^

This study aims to confirm these results in a multi-center double-blind, placebo controlled, parallel group study to evaluate the efficacy, safety and tolerability of an anthocyanin rich extract (ACRE) in subjects with moderately active UC.

## Materials and Methods

### Study Population

Patients with moderately or severely active UC were recruited between April 2019 and March 2021 at six IBD centers in Switzerland. Moderately or severely active UC was defined as a Mayo score 6-12 with an endoscopic sub-score ≥ 2. Patients aged 18–70 years and diagnosed with UC since at least three months with the disease extending at least 15 cm from the anal verge were included. Current oral or rectal 5-ASA/ sulfapyridine (SP) use or a history of oral or rectal 5-ASA/SP was allowed. Furthermore, patients were eligible for the study if they fulfilled one of the following criteria: a. Steroid intake up to 30 mg/day as well as a history of steroids dependency, refractory, or intolerance, including no steroids treatment due to earlier side-effects. OR b. Active disease despite induction therapy with 5-ASA agents, either mesalamine (2–4.8 g/day) or sulfasalazine (4–6 g/day) administered for at least two weeks. Topical treatment with 5-ASA was permissible but not sufficient for inclusion in the study. OR c. Intolerance to oral 5-ASA or azathioprine. OR d. Active disease despite thiopurine (adequately dosed according to treatment guidelines,^42^ such as 2-3 mg/kg for azathioprine) or methotrexate treatment administered for at least 12 weeks. OR e. Active disease despite treatment with biologics effective in UC or calcineurin inhibitors. No restrictions regarding other IBD therapies applied: Azathioprine/6-mercaptopurine was allowed, providing that the dose had been stable for 8 weeks prior to baseline and had been initiated at least two months before screening. TNF inhibitors (i.e. infliximab, adalimumab or golimumab) were allowed, providing that the dose was unchanged for at least two months prior to baseline and during the study treatment period. Vedolizumab and tofacitinib were allowed, providing that the dose remained unchanged for at least two months prior to baseline and during the study treatment period.

Exclusion criteria were a suspicion or diagnosis of Crohn’s disease, ischemic colitis, radiation colitis, indeterminate colitis, infectious colitis, diverticular disease associated colitis, microscopic colitis, massive pseudopolyposis or a colonic stenosis that could not be passed endoscopically, acute severe UC (as defined by Truelove and Witt’s criteria^43^) and/or signs of systemic toxicity, UC limited to the rectum (disease which extends <15 cm above the anal verge), long term treatment with antibiotics or non-steroidal anti-inflammatory drugs (NSAID) within two weeks prior to screening (one short treatment regime for antibiotics and occasional use of NSAID was allowed), and within at least 30 days after last treatment of the experimental product prior to enrolment history of malignancy, a history or presence of any clinically significant disorder that, in the opinion of the investigator, could impact on the patient’s ability to adhere to the study protocol and study procedures or would confound the study result or compromise patient safety.

All patients signed an informed consent form. The study was approved by the ethics committee of each center (BASEC2017-00156) and was conducted in accordance with the latest revision of Declaration of Helsinki^44^ as well as the guidelines of Good Clinical Practice.^45^ Moreover, the study was registered in the US online database of clinical research studies ClinicalTrials.gov (NCT04000139). All authors were offered full access to the study data and reviewed and approved the final manuscript.

### Study design and procedures

This was a multicenter randomized, double-blind, placebo-controlled trial. Patients with active moderate-severe UC despite state of the art 5—ASA, steroid, immunosuppressive of biological treatment were enrolled. Patients were randomized 2:1 (ACRE: placebo).

Upon study entry, all patients were instructed to continue their current medications unchanged. They either received a standardized anthocyanin extract (ACRE, referring to 800-1000 mg AC) administered *per os* three times daily or an optically identical placebo. Of note, similar or even lower AC doses have been previously administered in human studies investigating (extra)intestinal beneficial actions and with favorable results.^40,46,47^ As the ACRE capsule preparation contained purple compounds (which could not be eliminated during the production process), we added equally purple powder to the placebo capsule preparation, in order to avoid any unblinding. The total duration of investigational product administration amounted to eight weeks (56 days). All participating physicians were blinded to treatment assignment throughout the study. After enrollment, patients underwent a physical examination and laboratory blood tests including a complete blood count, liver function tests, and C-reactive protein, which were performed at both baseline and at the end of the phase protocol. Patients also underwent flexible rectosigmoidoscopy at study entry (baseline) and at week 8, and endoscopic activity was determined according to the endoscopic Mayo index sub-score^48^ by a local and a central reader.

### Clinical Assessment and Trial End Points

Primary objective of the study was to evaluate the efficacy of the ACRE preparation in subjects with moderately-severely active UC according to clinical, endoscopic, histologic and biochemical markers. Secondary objectives included the effects of ACRE on quality of life (QoL) as well as its safety.

(Mayo score 6-12, endoscopic sub-score ≥ 2) by comparing the clinical response rate of subjects on an ACRE versus a placebo arm at week 8. As clinical response was defined the reduction of total Mayo score (TMS) ≥ 3 points. Of note, the mentioned cut-off for clinical response exhibits an established validity in several studies previously.^49–51^ Secondary objectives were the evaluation of 1) e ACRE-efficacy at week 8 where clinical remission is defined as Mayo Score ≤ 2, with no individual sub-score > 1; 2) ACRE safety and tolerability; 3) efficacy of an ACRE preparation in subjects with moderately active UC compared to placebo in clinical remission, clinical response and clinical symptoms; 4) ACRE efficacy in subjects with moderately-severe active UC compared to placebo in endoscopic and histological remission and response; 5) quality of life (QOL) in patients of ACRE arm.

The primary endpoint was the clinical response at week 8 with clinical response defined as a reduction of TMS by ≥ 3 points, similar to previous studies.^49–51^

Secondary endpoints were:

- The proportion of patients with symptomatic remission at week 8, defined by the presence of both, a Mayo rectal bleeding sub-score of 0, and a stool frequency subscore of 0 or 1 (with at least one point decrease from baseline, week 0), (patient reported outcome) [PRO2].^52^
- The proportion of patients without rectal bleeding at week 8, defined by a Mayo rectal sub-score bleeding of 0.
- The proportion of patients with normal or enhanced stool frequency at week 8, defined by the Mayo stool frequency sub-score of 0 or 1 (with at least one point decrease from baseline, week 0).

Other secondary endpoints assessed rectal bleeding, clinical response and remission at week 4, durable remission (i.e. remission in weeks 8 and 12) and clinical response at week 8 according to the modified Mayo-score, defined as a three point and ≥30% drop from baseline of the sum of the rectal bleeding, stool frequency, endoscopy score (excluding friability) and physicians’ global assessment (PGA).

- The proportion of patients with endoscopic remission at week 8, defined by a modified Mayo endoscopic sub-score^48^ of 0 or 1 (excluding friability).
- The proportion of patients with histological remission at week 8, defined by a Geboes Index^53^ of grade 0 or 1.
- Mean change in fecal calprotectin concentrations at weeks 1, 2, 4, and 8 compared to baseline, week 0.
- Mean change in steroid dosage compared to baseline for patients in remission at week 8 to 12.
- Mean change in each of the short inflammatory bowel disease questionnaire (SIBDQ)^54^ sub-domains at week 8 compared to baseline (week 0).

The rate and time point of premature study withdrawals between both arms was also explored in the data analysis.

Data acquisition and management were performed using OpenClinica Community Edition, Version 3.14, and an OpenClinica eCRF.

### Analysis plan

A sample size of 112 patients was initially planned, leading to 100 patients completing the study, assuming a drop-out rate of 12%, based on a frequentist power calculation. According to our previous data, a clinical response was assumed in the active treatment arm of 55% and in the placebo of 25%. For a 1:1 placebo vs. verum randomization ratio, a patient number of 41 per group would be sufficient to achieve statistical significance with a power of 80%, assuming an α-error of maximal 5%. For the planned 1:2 ratio the respective numbers were be 33 (placebo) and 66 (verum) patients for both the groups. The analysis of the primary endpoint was conducted on the FAS (full analysis set) and PPS (per-protocol set). Data analysis was performed using SAS version 9.4 (Windows x64 version). Missing singular data items were carried forward. This did not apply, of course, when entire visits were missed (p.ex., in the case of premature study termination).

## RESULTS

### Study Population

Out of 48 patients screened in a total of six Swiss trial centers, 34 (70.9%) were enrolled and randomized, whereas 14 (29.1%; 4 females and 10 males) were considered screening failures.

Five randomized ACRE and two placebo patients prematurely terminated the study before the scheduled end of treatment at visit 3 (week 8). One ACRE patient started prednisone (20 mg/day) treatment at visit 1 and was excluded from all analyses for violation of the protocol. Thus, 18 ACRE and eight placebo patients could be analyzed in the per-protocol-set (PPS).

At screening, the majority of patients (69.2%) were on concomitant steroid medication. 16.7% in the bilberry group and 25% in the placebo group received anti-TNF treatment. Patient baseline characteristics are enclosed in Table 1.

**Table 1:** Baseline characteristics.

|  | *ACRE* | *Placebo* | *All patients* |
| --- | --- | --- | --- |
| Gender (f/m) | 10/14 | 5/5 | 15/19 |
| All randomized patients |  |  |  |
| Gender (f/m) | 8/10 | 4/4 | 12/14 |
| Per protocol set |  |  |  |
| Total Mayo Score<br>(Mean value ± SD) at<br>Screening | $8.43 \pm 1.31$ | $8.50 \pm 1.90$ | |
| Steroids/no steroids | 14 (77.8%) | 4 (50.0%) | 18 (69.2%) |
|  | 4 (22.2%) | 4 (50.0%) | 8 (30.8%) |
| TNF tx/no TNF tx | 3 (16.7%) | 2 (25.0%) | 5 (19.2%) |
|  | 15 (83.3%) | 6 (75.0%) | 21 (80.8%) |

ACRE, anthocyanin rich extract; f/m, female/male; SD, standard deviation; TNF,

|  | *Treatment* | *p-value* |
| --- | --- | --- |

tumor necrosis factor; tx, treatment

### Efficacy Analysis

#### Primary endpoint (Clinical Response at Week 8)

Clinical response at week 8 was achieved in nine out of 18 ACRE patients and three out of eight (3/8) placebo patients (Figure 1, Table 2). This corresponds to an odds ratio (OR) for response of 1.667 in favor of ACRE arm, with a 90% confidence interval (CI) between 0.399 – 6.963 and a one-sided p = 0.278 in the logistic analysis and the primary endpoint was not met.

**Figure 1:**
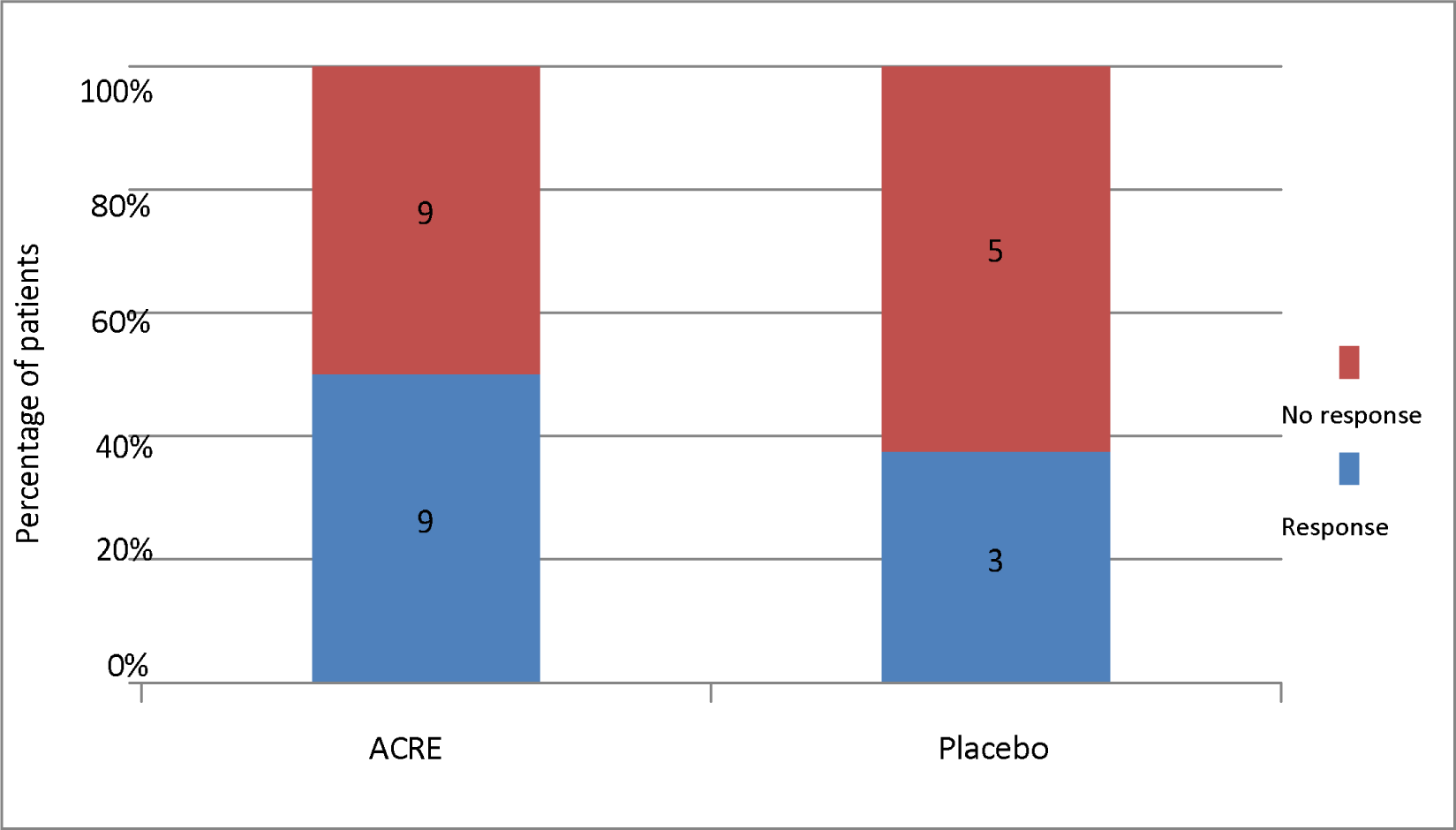
Clinical Response of both arms (ACRE vs placebo) at Week 8 after baseline. ACRE, Anthocyanin rich extract. Numbers within the bars indicate the absolute number of patients.

**Table 2:**
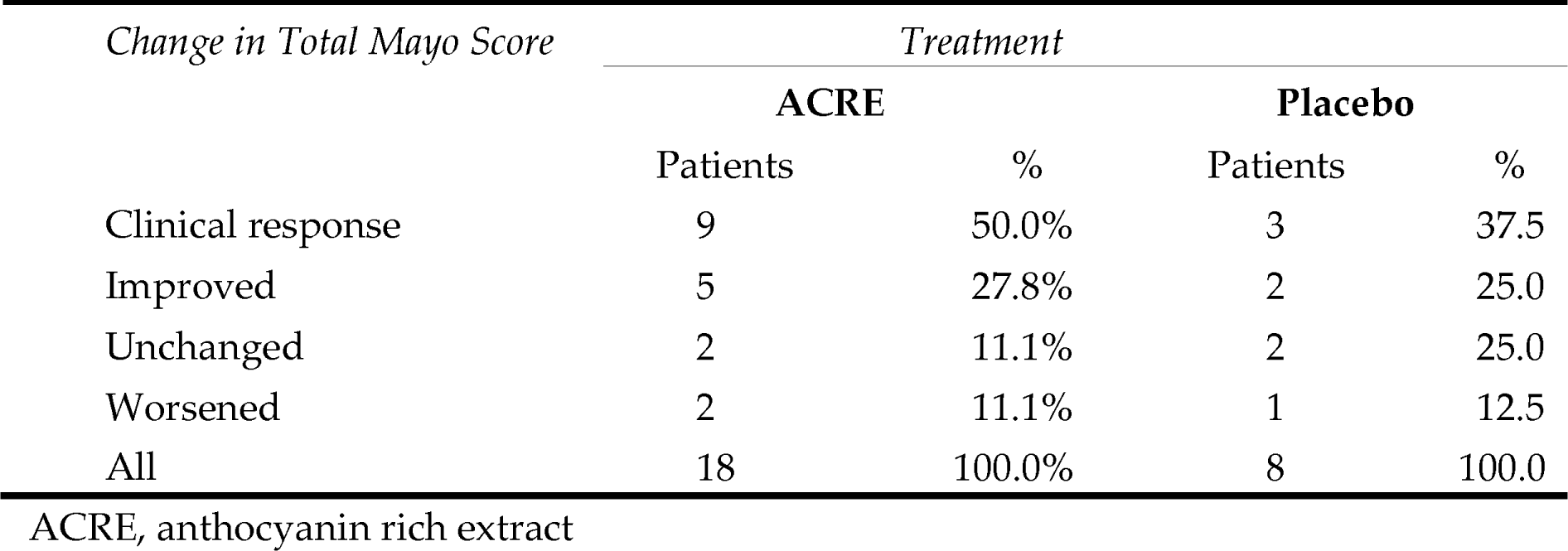
Categorized Change in Total Mayo Score.

| *Change in Total Mayo Score* | *Treatment* |  |  |  |
| --- | --- | --- | --- | --- |
|  | **ACRE** |  | **Placebo** |  |
|  | Patients | % | Patients | % |
| Clinical response | 9 | 50.0% | 3 | 37.5 |
| Improved | 5 | 27.8% | 2 | 25.0 |
| Unchanged | 2 | 11.1% | 2 | 25.0 |
| Worsened | 2 | 11.1% | 1 | 12.5 |
| All | 18 | 100.0% | 8 | 100.0 |

ACRE, anthocyanin rich extract

In a post-hoc analysis, mean reduction in partial Mayo score was 2.61 ± 2.79 for ACRE arm and 2.00 ± 3.07 for placebo arm (Figure 2, not significant). Assuming a normal distribution, the 90% CI of the change in TMS does not include the point 0 (no change) for ACRE while this point is within the 90% CI for placebo arm.

**Figure 2:**
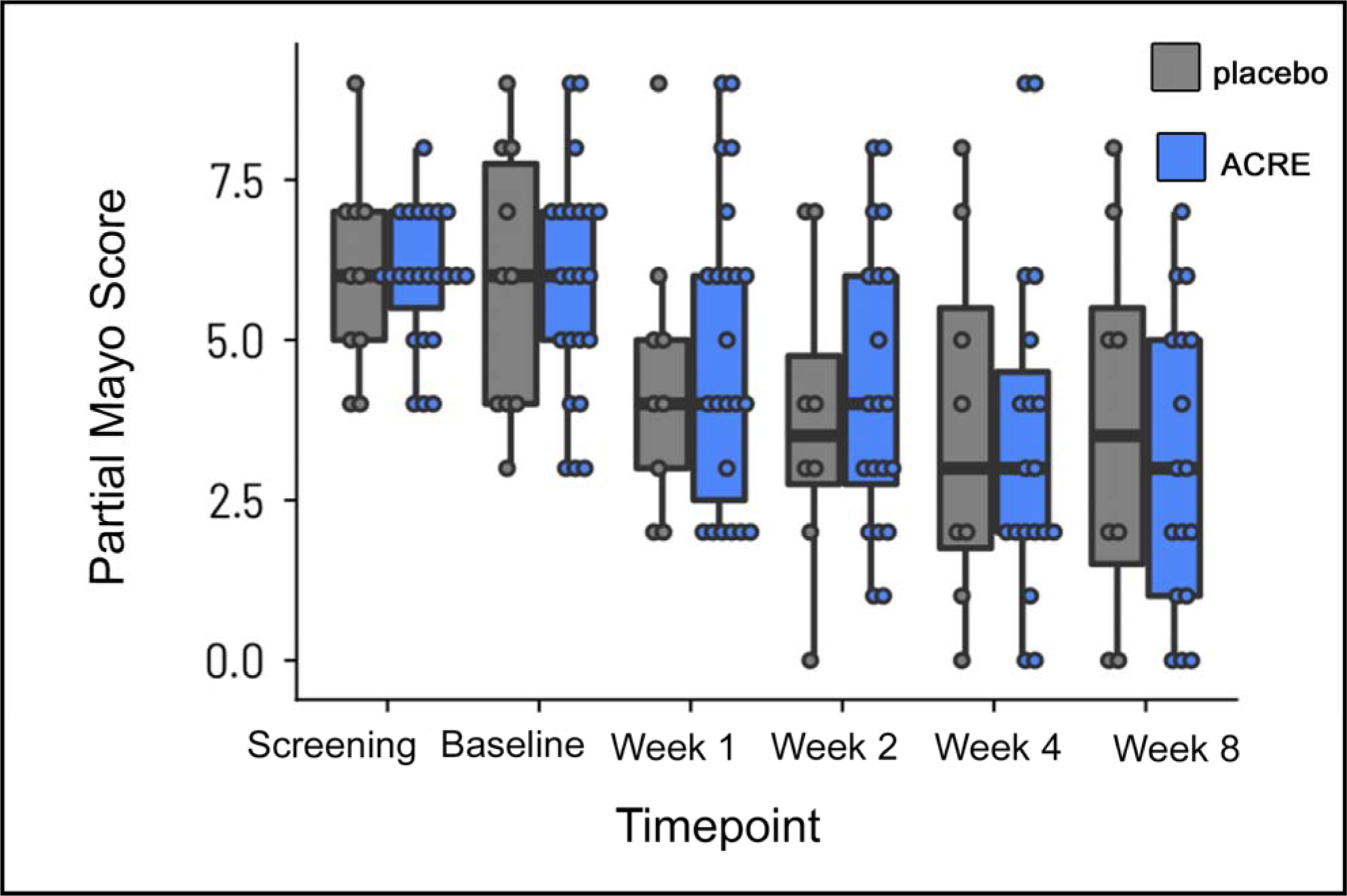
post-hoc analysis, mean reduction in partial Mayo score, with 90% confidence interval for mean, albeit without statistical significance. ACRE, anthocyanin rich extract.

**Figure 3:**
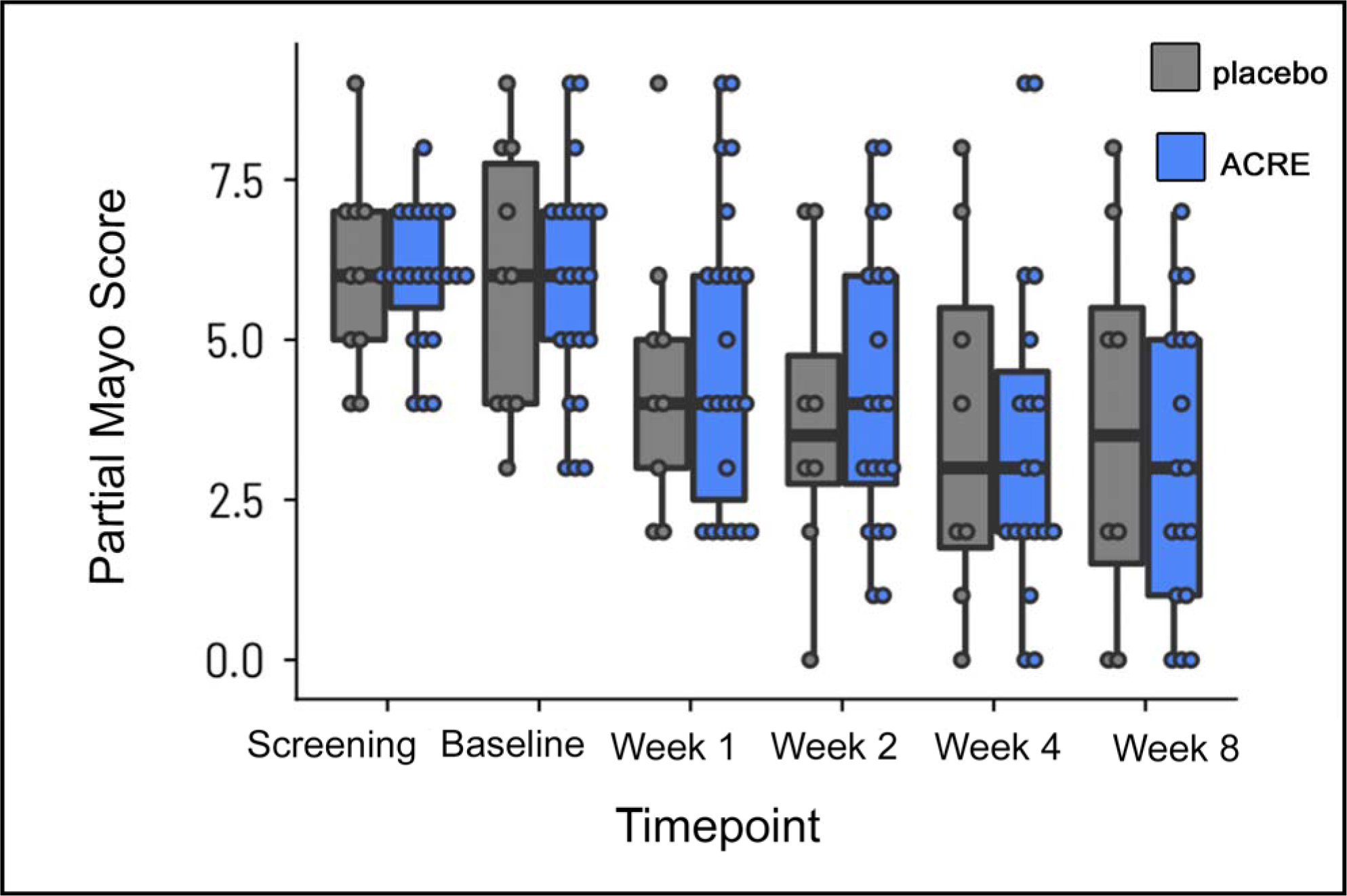
Calprotectin levels (μg/g) over the observation period between two arms. ACRE, anthocyanin rich extract.

An interesting finding of this study is the unusual high percentage of placebo response, i.e., 37.5% for the clinical response and 62.5% for the Mayo score amelioration.

#### Secondary endpoints

The secondary endpoint of clinical remission at week 8 was achieved in 9/18 patients of ACRE arm versus 3/8 patients of placebo arm (CI 0.399–6.963; p=0.278, not significant), even though we noted numerical improvements in both individual sub-scores rectal bleeding and stool frequency (Table 3).

**Table 3:** Categorized Changes at week 8.

|  | ACRE |  | Placebo |  |  |
| --- | --- | --- | --- | --- | --- |
|  | Patients | % | Patients | % |  |
| Symptomatic remission | 8/18 | 44.4 | 3/8 | 37.5 | 0.371 |
| No bleeding | 13/18 | 72.2 | 4/8 | 50.0 | 0.139 |
| Normal stool frequency | 10/18 | 11.1 | 4/8 | 50.0 | 0.397 |
| Endoscopic remission | 4/18 | 22.2 | 2/8 | 25 | 0.412 |

ACRE, anthocyanin rich extract

#### Endoscopic Remission at Week 8

The endoscopic findings (with central reading and site reading) yielded similar results, which are inconclusive and fail to demonstrate therapeutic effects.

#### Change in fecal calprotectin Endpoint vs. Baseline

Fecal calprotectin remained practically the same in placebo group between baseline and end of experiment values (947±1039 to 1040±1179) whereas it significantly decreased for ACRE group (1049±1139 to 557±756μg/g, p=0.035). No difference for any of the other secondary endpoints was noted.

#### Safety Analysis

Among the treated patients, a total of 68 adverse events, two of which were regarded to be serious, were recorded in 29 patients. All serious adverse events were assessed as unrelated to study medication. One patient developed an infectious colitis before enrolment. Another individual experienced an exacerbation of UC, which required hospitalization. Overall ACRE was very well tolerated and no new safety signals were detected.

## DISCUSSION

In this multi-center, randomized, double-blind and placebo controlled, phase IIa study, we aimed to evaluate the efficacy, safety as well as tolerability of ACRE in patients with UC. Although the clinical response at the end of the study did not differ between the two arms, a statistically significant difference was recorded in the secondary endpoint of fecal calprotectin.

Fecal calprotectin serves as a valuable indicator of inflammation within the intestines^55,56^ and has become an integral component of routine testing for diagnosing and monitoring IBD. According to a recent meta-analysis, fecal calprotectin is an inexpensive, valuable and rather accurate predictor IBD relapses.^55^ Additionally, further evidence has established fecal calprotectin as a useful marker recognizing an early response to IBD treatment, since it correlates robustly with serologic markers, endoscopic inflammation and disease activity indices of IBD subjects.^56^

As aforementioned, our research group firstly reported in 2013 within a pilot study^40^ the beneficial effects of bilberries on UC patients, results that we could also confirm later on.^35^ Comparable beneficial results of have been demonstrated independently by Kropat et al.^57^ An ongoing relevant Australian clinical trial with double-blind, randomized, controlled, multi-arm design has been recently announced,^58^ investigating the effect of administration of AC and/or multi-strain probiotics on UC patients.

In respect to the available *in vivo* preclinical scientific evidence, we reported as early as 2011 that bilberries and their AC positively impacted on an experimental acute and chronic colitis mouse model.^19^ Later, and again by utilizing the same murine model mimicking human UC and UC-associated cancer, we demonstrated^36^ that AC ingestion could avert the onset and progression of murine tumors in colon. Comparable favorable results of AC on IBD murine models have been henceforth published by other study groups.^29–31,59,60^

Interestingly, there is emerging evidence, that AC administration offers beneficial effects beyond IBD to further intestinal diseases such as colorectal carcinoma and irritable bowel syndrome.^61^ Bilberries and/or AC have also been investigated as a treatment targeting extraintestinal organs in various settings and have shown a positive impact in clinical trials, among others, for vascular health and cognitive function,^46^ or metabolic syndrome and associated conditions.^26^ A very recent US study with a large sample of patients (n = 37,232) found a direct inverse relationship between overall mortality risk and consumption of diverse berries and their containing flavonoids.^17^

One of the reasons for the health-promoting effects of AC is regarded to be their antioxidant effect owing to their phenolic structure, enabling them to neutralize ROS.^39^ Additionally, AC display antimicrobial effects, such as against *Bacillus cereus*, and *Helicobacter pylori.*^62^ They also reduce the expression of various genes involved in atherosclerosis formation in animal models.^63^ Moreover, AC inhibit inflammation-promoting pathways in immune cells.^39^

As stressed in the results section, an unusual high percentage of placebo response for the Mayo score improvement was noted. In this respect, a Cochrane meta-analysis^64^ of 61 studies estimated the placebo response and remission rates of induction treatment for UC (adult population) to be 33% and 12%, respectively. Placebo effect and rates of remission fluctuated based on several factors, including the severity of endoscopic disease and the rectal bleeding score upon trial initiation, the type of medication used, duration of the disease, and the specific time when the primary outcome was assessed.^64^

In a second more recent systematic review with meta-analysis (2022), clinical, endoscopic, histological and safety placebo rates in induction and maintenance trials of UC have been evaluated.^65^ After considering a total number of 119 trials (of which 92 in induction and 27 in maintenance) the abovementioned parameters were for the induction studies 11%, 19% and 15%, respectively. Again, the authors deduced placebo response rates observed in trials for UC differ depended on the endpoint evaluated, whether it pertains to response assessment or achieving remission, and whether the trial focuses on induction or maintenance.

The purple color of the drug as well as the placebo might be partially responsible for the high placebo rates. As a possible interpretation of this unexpected placebo effect, studies conducted in the past have identified the color of the placebo pills to correlate with outcomes.^66–68^ Thus, the colors of drugs may impact how their effects are perceived and may also play a role in their effectiveness. Additionally, there appears to be a connection between the coloring of drugs that affect the central nervous system and the conditions they are prescribed for. In this context there is evidence that red, yellow, and orange color of pills are linked to a stimulant effect, whereas blue, purple and green are connected to a tranquillizing – sedative effect.^66–68^

The strengths of this study includes the double-blinded, randomized and multi-center study design. The low number of participants is the main limitation of this study, this is at least partially due to restriction during the COVID-19 pandemic. since individual patients, as aforementioned commented, have impacted relevantly the outcome. However, this did not prevent a statistical significance for fecal calprotectin between the two study arms.

## CONCLUSION

In conclusion, ACRE did not induce clinical response at 8 weeks in patients with moderate to severe UC. However, due to limitations to recruit a large sample size, limiting some analyses, significant decreases in fecal calprotectin levels upon ACRE treatment were noted.

## CONFLICTS OF INTEREST

PS received consulting fees from Pfizer, Abbvie, Takeda and Janssen-Cilag and travel support from Falk, UCB and Pfizer.

LB reports fees for consulting/advisory board from Abbvie, MSD, Vifor, Falk, Esocap, Calypso, Ferring, Pfizer, Shire, Takeda, Janssen, Ewopharma.

GR declares consulting fees from Abbvie, Augurix, BMS, Boehringer, Calypso, Celgene, FALK, Ferring, Fisher, Genentech, Gilead, Janssen, MSD, Novartis, Pfizer, Phadia, Roche, UCB, Takeda, Tillots, Vifor, Vital Solutions and Zeller; speaker’s honoraria from Astra Zeneca, Abbvie, FALK, Janssen, MSD, Pfizer, Phadia, Takeda, Tillots, UCB, Vifor and Zeller; and grants support from Abbvie, Ardeypharm, Augurix, Calypso, FALK, Flamentera, MSD, Novartis, Pfizer, Roche, Takeda, Tillots, UCB and Zeller.

BM reports traveling fees from Takeda, Vifor, Gilead and MSD. BM received fees as a speaker from Takeda. BM has served at an advisory board for Abbvie, Gilead, Takeda and BMS. BM has received research grants from MSD, BMS and Nestlé unrelated to the submitted work.

MD reports traveling fees from Takeda, FALK, Abbvie as well as consulting fees from Takeda.

Rest of the authors declare no compelling conflict of interests.

## ACKNOWLEDGEMENTS

The authors wish to thank all the patients for their cooperation as well as their families for their support during this endeavor.

## Funding

This work was supported by grants from the Swiss National Science Foundation (SNF) to GR [Grant No. 33IC30_166844] and the Litwin Foundation (New Hyde Park, NY).

## Clinicaltrials.gov number

NCT04000139

## Ethical approval number

BASEC2017-00156

## Informed consent statement

All patients signed an informed consent form

## Data availability statement

Data available on request due to privacy/ethical restrictions

## Conflicts of interest

PS received consulting fees from Pfizer, Abbvie, Takeda and Janssen-Cilag and travel support from Falk, UCB and Pfizer.LB reports fees for consulting/advisory board from Abbvie, MSD, Vifor, Falk, Esocap, Calypso, Ferring, Pfizer, Shire, Takeda, Janssen, Ewopharma.

BM reports traveling fees from Takeda, Vifor, Gilead and MSD. BM received fees as a speaker from Takeda. BM has served at an advisory board for Gilead, Takeda and BMS. BM has received research grants from MSD and BMS unrelated to the submitted work.

MD reports traveling fees from Takeda, FALK, Abbvie as well as consulting fees from Takeda.

Rest of the authors declare no compelling conflict of interests.

## Abbreviations

5-ASA, aminosalicylic acid (mesalazine); AC, anthocyanins; ACRE, anthocyanin rich extract; CI, confidence interval; IBD, inflammatory bowel disease; IFN, interferon; IL, interleukin; NF-κB, nuclear factor kappa B; NSAID, non-steroidal anti-inflammatory drugs; OR, odds ratio; PPS, Per-Protocol-Set; ROS, reactive oxygen species; PRO, patient reported outcome; QOL, quality of life; SIBDQ, short inflammatory bowel disease questionnaire; TNF, tumor necrosis factor; TMS, total Mayo score; UC, ulcerative colitis

## Notes

### Clinical Trial

NCT04000139

### Funding Statement

This study was funded by the Swiss National Science Foundation (SNF) to GR [Grant No. 33IC30_166844] and the Litwin Foundation (New Hyde Park, NY)

### Author Declarations

Swiss ethics committee approval approval number: BASEC2017-00156, Date of authorisation by the ethics committee 14.02.2019 Authorisation initially for Canton Zurich (University Hospital, Prof. Rogler) and then extension of the approval to the rest participating centers (Basel, Bern, Geneva, Lausanne, St. Gallen)

## REFERENCES

1. Lang BM, Ledergerber M, Jordi SBU, et al. Because I’m happy - positive affect and its predictive value for future disease activity in patients with inflammatory bowel diseases: a retrospective cohort study. Therap Adv Gastroenterol. 2023;16:17562848231179335.

2. Ungaro R, Mehandru S, Allen PB, Peyrin-Biroulet L, Colombel JF. Ulcerative colitis. Lancet. 2017;389(10080):1756–1770.

3. Norouzkhani N, Faramarzi M, Ghodousi Moghadam S, et al. Identification of the informational and supportive needs of patients diagnosed with inflammatory bowel disease: a scoping review. Front Psychol. 2023;14:1055449.

4. Awadhi SA, Alboraie M, Albaba EA, et al. Treatment of Patients with Mild to Moderate Ulcerative Colitis: A Middle East Expert Consensus. J Clin Med. 2023;12(21).

5. Harbord M, Eliakim R, Bettenworth D, et al. Third European Evidence-based Consensus on Diagnosis and Management of Ulcerative Colitis. Part 2: Current Management. J Crohns Colitis. 2017;11(7):769-784.

6. Dai N, Haidar O, Askari A, Segal JP. Colectomy rates in ulcerative colitis: A systematic review and meta-analysis. Dig Liver Dis. 2023;55(1):13–20.

7. Wang K, Zhu Y, Liu K, Zhu H, Ouyang M. Adverse events of biologic or small molecule therapies in clinical trials for inflammatory bowel disease: A systematic review and meta-analysis. Heliyon. 2024;10(4):e25357.

8. Stallmach A, Hagel S, Bruns T. Adverse effects of biologics used for treating IBD. Best Pract Res Clin Gastroenterol. 2010;24(2):167–182.

9. Larussa T, Basile A, Palleria C, et al. Real-life burden of adverse reactions to biological therapy in inflammatory bowel disease: a single-centre prospective case series. Med Pharm Rep. 2021;94(3):289–297.

10. Alulis S, Vadstrup K, Olsen J, et al. The cost burden of Crohn’s disease and ulcerative colitis depending on biologic treatment status - a Danish register-based study. BMC Health Serv Res. 2021;21(1):836.

11. van Linschoten RCA, Visser E, Niehot CD, et al. Systematic review: societal cost of illness of inflammatory bowel disease is increasing due to biologics and varies between continents. Aliment Pharmacol Ther. 2021;54(3):234–248.

12. Lasa JS, Olivera PA, Danese S, Peyrin-Biroulet L. Efficacy and safety of biologics and small molecule drugs for patients with moderate-to-severe ulcerative colitis: a systematic review and network meta-analysis. Lancet Gastroenterol Hepatol. 2022;7(2):161–170.

13. Pituch-Zdanowska A, Dembinski L, Banaszkiewicz A. Old but Fancy: Curcumin in Ulcerative Colitis-Current Overview. Nutrients. 2022;14(24).

14. Gupta M, Mishra V, Gulati M, et al. Natural compounds as safe therapeutic options for ulcerative colitis. Inflammopharmacology. 2022;30(2):397–434.

15. Triantafyllidi A, Xanthos T, Papalois A, Triantafillidis JK. Herbal and plant therapy in patients with inflammatory bowel disease. Ann Gastroenterol. 2015;28(2):210–220.

16. Lang Y, Gao N, Zang Z, et al. Classification and antioxidant assays of polyphenols: a review. Journal of Future Foods. 2024;4(3):193–204.

17. Zhang L, Muscat JE, Chinchilli VM, Kris-Etherton PM, Al-Shaar L, Richie JP. Consumption of Berries and Flavonoids in Relation to Mortality in NHANES, 1999-2014. J Nutr. 2024;154(2):734-743.

18. Krga I, Milenkovic D. Anthocyanins: From Sources and Bioavailability to Cardiovascular-Health Benefits and Molecular Mechanisms of Action. J Agric Food Chem. 2019;67(7):1771–1783.

19. Piberger H, Oehme A, Hofmann C, et al. Bilberries and their anthocyanins ameliorate experimental colitis. Mol Nutr Food Res. 2011;55(11):1724–1729.

20. Tena N, Martin J, Asuero AG. State of the Art of Anthocyanins: Antioxidant Activity, Sources, Bioavailability, and Therapeutic Effect in Human Health. Antioxidants (Basel). 2020;9(5).

21. Ahnfelt-Ronne I, Nielsen OH, Christensen A, Langholz E, Binder V, Riis P. Clinical evidence supporting the radical scavenger mechanism of 5-aminosalicylic acid. Gastroenterology. 1990;98(5 Pt 1):1162-1169.

22. Nielsen OH, Bukhave K, Elmgreen J, Ahnfelt-Ronne I. Inhibition of 5-lipoxygenase pathway of arachidonic acid metabolism in human neutrophils by sulfasalazine and 5-aminosalicylic acid. Dig Dis Sci. 1987;32(6):577–582.

23. Allgayer H, Stenson WF. A comparison of effects of sulfasalazine and its metabolites on the metabolism of endogenous vs. exogenous arachidonic acid. Immunopharmacology. 1988;15(1):39–46.

24. Al-Khayri JM, Sahana GR, Nagella P, Joseph BV, Alessa FM, Al-Mssallem MQ. Flavonoids as Potential Anti-Inflammatory Molecules: A Review. Molecules. 2022;27(9).

25. Sahoo DK, Heilmann RM, Paital B, et al. Oxidative stress, hormones, and effects of natural antioxidants on intestinal inflammation in inflammatory bowel disease. Front Endocrinol (Lausanne). 2023;14:1217165.

26. Sharma A, Lee HJ. Anti-Inflammatory Activity of Bilberry (Vaccinium myrtillus L.). Curr Issues Mol Biol. 2022;44(10):4570–4583.

27. Akiyama S, Nesumi A, Maeda-Yamamoto M, Uehara M, Murakami A. Effects of anthocyanin-rich tea “Sunrouge” on dextran sodium sulfate-induced colitis in mice. Biofactors. 2012;38(3):226–233.

28. Li L, Wang L, Wu Z, et al. Anthocyanin-rich fractions from red raspberries attenuate inflammation in both RAW264.7 macrophages and a mouse model of colitis. Sci Rep. 2014;4:6234.

29. Zhao L, Zhang Y, Liu G, Hao S, Wang C, Wang Y. Black rice anthocyanin-rich extract and rosmarinic acid, alone and in combination, protect against DSS-induced colitis in mice. Food Funct. 2018;9(5):2796–2808.

30. Peng Y, Yan Y, Wan P, et al. Gut microbiota modulation and anti-inflammatory properties of anthocyanins from the fruits of Lycium ruthenicum Murray in dextran sodium sulfate-induced colitis in mice. Free Radic Biol Med. 2019;136:96–108.

31. Li S, Wang T, Wu B, et al. Anthocyanin-containing purple potatoes ameliorate DSS-induced colitis in mice. J Nutr Biochem. 2021;93:108616.

32. Mu J, Xu J, Wang L, Chen C, Chen P. Anti-inflammatory effects of purple sweet potato anthocyanin extract in DSS-induced colitis: modulation of commensal bacteria and attenuated bacterial intestinal infection. Food Funct. 2021;12(22):11503–11514.

33. Zhang G, Gu Y, Dai X. Protective Effect of Bilberry Anthocyanin Extracts on Dextran Sulfate Sodium-Induced Intestinal Damage in Drosophila melanogaster. Nutrients. 2022;14(14).

34. Roth S, Spalinger MR, Muller I, Lang S, Rogler G, Scharl M. Bilberry-derived anthocyanins prevent IFN-gamma-induced pro-inflammatory signalling and cytokine secretion in human THP-1 monocytic cells. Digestion. 2014;90(3):179–189.

35. Roth S, Spalinger MR, Gottier C, et al. Bilberry-Derived Anthocyanins Modulate Cytokine Expression in the Intestine of Patients with Ulcerative Colitis. PLoS One. 2016;11(5):e0154817.

36. Lippert E, Ruemmele P, Obermeier F, et al. Anthocyanins Prevent Colorectal Cancer Development in a Mouse Model. Digestion. 2017;95(4):275–280.

37. Dreiseitel A, Schreier P, Oehme A, et al. Anthocyanins and anthocyanidins are poor inhibitors of CYP2D6. Methods Find Exp Clin Pharmacol. 2009;31(1):3–9.

38. Dreiseitel A, Schreier P, Oehme A, et al. Inhibition of proteasome activity by anthocyanins and anthocyanidins. Biochem Biophys Res Commun. 2008;372(1):57–61.

39. Scharl M, Rogler G, Biedermann L. Anthocyane, Heidelbeeren und Curcuma: Wirksame Therapeutika bei Darmentzündungen? Schweizerische Zeitschrift für Ganzheitsmedizin / Swiss Journal of Integrative Medicine. 2017;29(3):137–140.

40. Biedermann L, Mwinyi J, Scharl M, et al. Bilberry ingestion improves disease activity in mild to moderate ulcerative colitis - an open pilot study. J Crohns Colitis. 2013;7(4):271–279.

41. Li S, Wu B, Fu W, Reddivari L. The Anti-inflammatory Effects of Dietary Anthocyanins against Ulcerative Colitis. Int J Mol Sci. 2019;20(10).

42. Rubin DT, Ananthakrishnan AN, Siegel CA, Sauer BG, Long MD. ACG Clinical Guideline: Ulcerative Colitis in Adults. Am J Gastroenterol. 2019;114(3):384–413.

43. Truelove SC, Witts LJ. Cortisone in ulcerative colitis; final report on a therapeutic trial. Br Med J. 1955;2(4947):1041-1048.

44. World Medical A. World Medical Association Declaration of Helsinki: ethical principles for medical research involving human subjects. JAMA. 2013;310(20):2191–2194.

45. (R3) IE. Guideline on good clinical practice (GCP) Step 2b. 2023; https://www.ema.europa.eu/en/ich-e6-r2-good-clinical-practice-scientific-guideline. Accessed 15.03.2024.

46. Wood E, Hein S, Mesnage R, et al. Wild blueberry (poly)phenols can improve vascular function and cognitive performance in healthy older individuals: a double-blind randomized controlled trial. Am J Clin Nutr. 2023;117(6):1306–1319.

47. Zhang H, Xu Z, Zhao H, et al. Anthocyanin supplementation improves anti-oxidative and anti-inflammatory capacity in a dose-response manner in subjects with dyslipidemia. Redox Biol. 2020;32:101474.

48. Schroeder KW, Tremaine WJ, Ilstrup DM. Coated oral 5-aminosalicylic acid therapy for mildly to moderately active ulcerative colitis. A randomized study. N Engl J Med. 1987;317(26):1625–1629.

49. Lewis JD, Chuai S, Nessel L, Lichtenstein GR, Aberra FN, Ellenberg JH. Use of the noninvasive components of the Mayo score to assess clinical response in ulcerative colitis. Inflamm Bowel Dis. 2008;14(12):1660–1666.

50. Sandborn WJ, Sands BE, Vermeire S, et al. Modified Mayo score versus Mayo score for evaluation of treatment efficacy in patients with ulcerative colitis: data from the tofacitinib OCTAVE program. Therap Adv Gastroenterol. 2022;15:17562848221136331.

51. Sandborn WJ, Cyrille M, Hansen MB, et al. Efficacy and Safety of Abrilumab in a Randomized, Placebo-Controlled Trial for Moderate-to-Severe Ulcerative Colitis. Gastroenterology. 2019;156(4):946–957 e918.

52. Bewtra M, Brensinger CM, Tomov VT, et al. An optimized patient-reported ulcerative colitis disease activity measure derived from the Mayo score and the simple clinical colitis activity index. Inflamm Bowel Dis. 2014;20(6):1070–1078.

53. Geboes K, Riddell R, Ost A, Jensfelt B, Persson T, Lofberg R. A reproducible grading scale for histological assessment of inflammation in ulcerative colitis. Gut. 2000;47(3):404–409.

54. Irvine EJ, Zhou Q, Thompson AK. The Short Inflammatory Bowel Disease Questionnaire: a quality of life instrument for community physicians managing inflammatory bowel disease. CCRPT Investigators. Canadian Crohn’s Relapse Prevention Trial. Am J Gastroenterol. 1996;91(8):1571–1578.

55. Shi JT, Chen N, Xu J, et al. Diagnostic Accuracy of Fecal Calprotectin for Predicting Relapse in Inflammatory Bowel Disease: A Meta-Analysis. J Clin Med. 2023;12(3).

56. Lee YW, Lee KM, Lee JM, et al. The usefulness of fecal calprotectin in assessing inflammatory bowel disease activity. Korean J Intern Med. 2019;34(1):72–80.

57. Kropat C, Mueller D, Boettler U, et al. Modulation of Nrf2-dependent gene transcription by bilberry anthocyanins in vivo. Mol Nutr Food Res. 2013;57(3):545–550.

58. Cosier D, Lambert K, Batterham M, Sanderson-Smith M, Mansfield KJ, Charlton K. The INHABIT (synergIstic effect of aNtHocyAnin and proBIoTics in) Inflammatory Bowel Disease trial: a study protocol for a double-blind, randomised, controlled, multi-arm trial. J Nutr Sci. 2024;13:e1.

59. Chen J, Jiang F, Xu N, et al. Anthocyanin Extracted from Purple Sweet Potato Alleviates Dextran Sulfate Sodium-Induced Colitis in Mice by Suppressing Pyroptosis and Altering Intestinal Flora Structure. J Med Food. 2024;27(2):110–122.

60. Mo J, Ni J, Zhang M, et al. Mulberry Anthocyanins Ameliorate DSS-Induced Ulcerative Colitis by Improving Intestinal Barrier Function and Modulating Gut Microbiota. Antioxidants (Basel). 2022;11(9).

61. Liu Y, Fernandes I, Mateus N, Oliveira H, Han F. The Role of Anthocyanins in Alleviating Intestinal Diseases: A Mini Review. J Agric Food Chem. 2024.

62. Nohynek LJ, Alakomi HL, Kahkonen MP, et al. Berry phenolics: antimicrobial properties and mechanisms of action against severe human pathogens. Nutr Cancer. 2006;54(1):18–32.

63. Mauray A, Felgines C, Morand C, Mazur A, Scalbert A, Milenkovic D. Bilberry anthocyanin-rich extract alters expression of genes related to atherosclerosis development in aorta of apo E-deficient mice. Nutr Metab Cardiovasc Dis. 2012;22(1):72–80.

64. Jairath V, Zou GY, Parker CE, et al. Placebo response and remission rates in randomised trials of induction and maintenance therapy for ulcerative colitis. Cochrane Database Syst Rev. 2017;9(9):CD011572.

65. Sedano R, Hogan M, Nguyen TM, et al. Systematic Review and Meta-Analysis: Clinical, Endoscopic, Histological and Safety Placebo Rates in Induction and Maintenance Trials of Ulcerative Colitis. J Crohns Colitis. 2022;16(2):224–243.

66. Bhugra D, Ventriglio A, Till A, Malhi G. Colour, culture and placebo response. Int J Soc Psychiatry. 2015;61(6):615–617.

67. de Craen AJ, Roos PJ, de Vries AL, Kleijnen J. Effect of colour of drugs: systematic review of perceived effect of drugs and of their effectiveness. BMJ. 1996;313(7072):1624-1626.

68. Jacobs KW, Nordan FM. Classification of placebo drugs: effect of color. Percept Mot Skills. 1979;49(2):367–372.

